# Angiotensin converting enzyme inhibitors and risk of lung cancer: a population-based cohort study

**DOI:** 10.1101/2021.01.04.21249217

**Authors:** Joshua Kai Fung Hung, Jiandong Zhou, Sharen Lee, Yunlong Xia, Ying Liu, Yuhui Zhang, Wing Tak Wong, Guangling Li, Tong Liu, Bernard Man Yung Cheung, Ian Chi Kei Wong, Qingpeng Zhang, Gary Tse

## Abstract

**Objectives:** To determine whether the use of angiotensin converting enzyme inhibitors (ACEIs) was associated with a higher risk of lung cancer when compared to use of angiotensin receptor blockers (ARBs).

**Study Design:** Population-based cohort study.

**Setting:** Public hospitals under the Hospital Authority in Hong Kong, P.R. China.

**Methods:** Patients admitted to public hospitals and first prescribed with ACEI and/or ARB between 1 January 2001 and 31 December 2018 were analyzed. The last follow-up date was 31 August 2020, or death, whichever was earlier.

**Outcomes:** The primary outcome was the incidence of lung cancer. Logistic regression was used to calculate odds ratio [ORs] with 95% confidence intervals associated with the use of ACEIs compared to ARBs. Incidence and odds ratios were estimated for temporal analysis of incident cancer risk associated with time since the first prescription of ACEI or ARB.

**Results:** In the unmatched cohort, 56,697 patients and 357,011 patients were included the ARB and ACEI cohorts, with lung cancer incidence of 2.16% and 1.29%, respectively. Using 1:3 matching for ARB to ACEI users, the incidences were 2.32% and 1.29%. ACEI use was associated with increased risks of lung cancer both before (hazard ratio: 1.30 [1.21-1.40], P<0.0001) and after (1.40 [1.29-1.51], P<0.0001) matching. There was a dose-dependent relationship between ACEI exposure and lung cancer risk.

**Conclusions:** ACEI use was associated with increased risk of lung cancer compared with ARB use at all time points. Additionally, incidence risk increases with the duration of exposure.

## Introduction

The global disease burden of hypertension has long been recognized. Recent studies using more than 10 million death records from the Centers for Disease Control and Prevention database reported a significant increase in the age-related hypertension-related cardiovascular death rate from 18.3 to 23.0 between 2007 and 2017 in the United States [1]. Approximately half of the adult population in the United States are estimated to suffer from hypertension [2]. Angiotensin-converting enzyme inhibitors (ACEIs) are the first-line agents effective in the treatment of hypertension. Angiotensin is a hormone involved in the renin-angiotensin system responsible for systemic autoregulation of blood pressure and fluid-electrolyte balance. Low blood volume or hyponatremia results in reduced renal blood flow, triggering the release of renin from juxtaglomerular cells into the circulation. ACEIs prevents the conversion of angiotensin I to angiotensin II, whereas angiotensin receptor blockers (ARBs) act further downstream by blocking the receptors of angiotensin. Together, these reduce the vasoconstrictive effects of angiotensin II and the subsequent release of mineralocorticoids, thereby lowering blood pressure.

Although commonly prescribed, the use of ACEIs has been associated with adverse effects. A persistent dry cough following ACEI initiation is commonly reported, with patients of Chinese descent more likely to discontinue medication use due to ACEI-induced cough [3]. ACE is responsible for the breakdown of bradykinin and substance P, which act as mediators of cough in the upper airway and lungs [4]. Moreover, both bradykinin [5] and substance P were found to be associated with oncogenic effects [6]. Therefore, ACEI may be associated with cancer risk. A recent population-based cohort study in the United Kingdom reported a 14% increase in lung cancer incidence with long-term use of ACEIs compared to ARBs [7]. Similarly, ACEIs are also reported to have higher lung cancer risks compared to ARBs in Asians [8]. However, systematic reviews and meta-analyses have found no increase in cancer risks from pooled data of randomized controlled trials [9, 10]. Moreover, other than the lungs, organ systems such as the gastrointestinal tract also show ACE expression. In this exploratory study, we tested the hypothesis that ACEI users have higher lung and gastrointestinal cancer risks using population-based healthcare data from Hong Kong.

## Methods

### Study Design and Data Sources

Ethics approval for the present study was obtained from The Joint Chinese University of Hong Kong-New Territories East Cluster Clinical Research Ethics Committee. The present retrospective cohort study obtained its data from the Hong Kong Clinical Data Analysis and Reporting System (CDARS). CDARS is a territory-wide healthcare database that centralizes information from 42 public hospitals, 48 specialist outpatient clinics, and 73 general outpatient clinics, serving nearly 7.5 million people. The database contains electrical health records including demographic information and all clinical diagnoses (coded with ICD 9 classification). All patient information, drug prescription details, and laboratory investigation from inpatient, outpatient, and accident and emergency admissions are available.

### Study Population and Bias Reduction

Patients aged 18 or older admitted to public hospitals under the Hong Kong Hospital Authority, and prescribed ACEIs or ARB between 1 January 2001 and 31 December 2017 were identified. This includes patients prescribed anti-hypertensive medication and those switching to ACEI/ARB from other anti-hypertensive medication classes. Patients were categorized into two study cohorts: an ACEI group and an ARB group.

The patients were identified from the Clinical Data Analysis and Reporting System (CDARS), a territory-wide database that centralizes patient information from individual local hospitals to establish comprehensive medical data, including demographics, comorbidities, and ACEI/ARB and other drug prescriptions (**Table 1** and **Table 2**). The system has been previously used by both our team and other teams in Hong Kong. The population of interest has prior comorbidities including cardiovascular diseases, respiratory system diseases, renal diseases, endocrine diseases, diabetes mellitus, hypertension, gastroenterology, and obesity, before first prescription date of ACEI/ARB.

**Table 1.** Clinical baseline characteristics of the unmatched cohort. * for p≤ 0.05, ** for p ≤ 0.01, *** for p ≤ 0.001

| Characteristics | ARB (N=56697) | ACEI (N=357011) | P value |
| --- | --- | --- | --- |
|  | Median (IQR);Max;N or Count(%) | Median (IQR);Max;N or Count(%) |  |
| Mortality | 7785(13.73%) | 187962(52.64%) | <0.0001*** |
| Duration to mortality, days | 1665(1074-2742);7012;n=56697 | 2828(1366-4403);7087;n=357011 | <0.0001*** |
| Lung cancer | 733(1.29%) | 7719(2.16%) | <0.0001*** |
| Duration to lung cancer, days | 1646.0(1060-2717);7104;n=56697 | 2814(1353-4395);7087;n=357011 | <0.0001*** |
| **Demographics** |  |  |  |
| Male gender | 25995(45.84%) | 193387(54.16%) | <0.0001*** |
| Baseline age, year | 70.6(60.1-80.1);110.2;n=56697 | 71.60-79.2854);116.3;n=357011 | 0.1612 |
| **Past comorbidities** |  |  |  |
| Cardiovascular | 22946(40.47%) | 143090(40.07%) | 0.2507 |
| Respiratory | 17188(30.31%) | 108415(30.36%) | 0.8589 |
| Renal | 12007(21.17%) | 75182(21.05%) | 0.6070 |
| Endocrine | 725(1.27%) | 3237(0.90%) | <0.0001*** |
| Diabetes mellitus | 8935(15.75%) | 63840(17.88%) | <0.0001*** |
| Hypertension | 32359(57.07%) | 175030(49.02%) | <0.0001*** |
| Gastrointestinal | 20308(35.81%) | 114068(31.95%) | <0.0001*** |
| **Medications** |  |  |  |
| ACEI use | 0(0.00%) | 357011(100.00%) | <0.0001*** |
| ACEI dosage, $\times 10^3$ mg | - | 5.92(0.86-24.5);5234.9;n=357010 | - |
| ACEI duration, days | - | 1273(217-3246);7066;n=306269 | - |
| ARB use | 56697(100.00%) | 0(0.00%) | <0.0001*** |
| ARB dosage, x10<sup>4</sup> mg | 7.8(1.98-19.2);427.32;n=8065 | - | - |
| ARB duration, days | 1694(821-2885);6870;n=7796 | - | - |
| Beta blockers | 26315(46.41%) | 164812(46.16%) | 0.5056 |
| Calcium channel blockers | 34413(60.69%) | 200581(56.18%) | <0.0001*** |
| Diuretics | 19646(34.65%) | 144852(40.57%) | <0.0001*** |

**Table 2.** Baseline characteristics of the matched cohort. * for p≤ 0.05, ** for p ≤ 0.01, *** for p ≤ 0.001

| Characteristics | ARB (N=56697) | ACEI (N=170091) | P value |
| --- | --- | --- | --- |
|  | Median (IQR);Max;N or Count(%) | Median (IQR);Max;N or Count(%) |  |
| Mortality | 7785(13.73%) | 85135(50.05%) | <0.0001*** |
| Duration to mortality, days | 1665(1074-2742);7026;n=56697 | 2923(1452-4498);7100;n=170091 | <0.0001*** |
| Lung cancer | 733(1.29%) | 3951(2.32%) | <0.0001*** |
| Duration to lung cancer, days | 1646(1060-2717);7012;n=56697 | 2911(1436-4488);7018;n=170091 | <0.0001*** |
| **Demographics** |  |  |  |
| Male gender | 25995(45.84%) | 78006(45.86%) | 0.9786 |
| Baseline age, year | 70.6(60.1-80.1);110.2;n=56697 | 70.6(60.2-80.1);105;n=170091 | 0.9745 |
| **Past comorbidities** |  |  |  |
| Cardiovascular | 22946(40.47%) | 68838(40.47%) | 1.0000 |
| Respiratory | 17188(30.31%) | 51566(30.31%) | 0.9991 |
| Renal | 12007(21.17%) | 36020(21.17%) | 0.9973 |
| Endocrine | 725(1.27%) | 2175(1.27%) | 1.0000 |
| Diabetes mellitus | 8935(15.75%) | 26805(15.75%) | 1.0000 |
| Hypertension | 32359(57.07%) | 97145(57.11%) | 0.9338 |
| Gastrointestinal | 20308(35.81%) | 60924(35.81%) | 1.0000 |
| **Medications** |  |  |  |
| ACEI use | 0(0.00%) | 170091(100.00%) | <0.0001*** |
| ACEI dosage, x10<sup>3</sup> mg | - | 6.2 (0.9-25.3);271.2;n=170090 | - |
| ACEI duration, days | - | 1298(228-3306);4408;n=146863 | - |
| ARB use | 56697(100.00%) | 0(0.00%) | <0.0001*** |
| ARB dosage, x10<sup>4</sup> mg | 7.84 (1.98-19.2);127.3;n=8065 | - | - |
| ARB duration, days | 1694(821-2885);5870;n=7796 | - | - |
| Beta blockers | 26315(46.41%) | 78847(46.35%) | 0.8887 |
| Calcium channel blockers | 34413(60.69%) | 103244(60.69%) | 0.9983 |
| Diuretics | 19646(34.65%) | 58934(34.64%) | 0.9981 |

Drug prescription details (cumulative dosage and cumulative duration) of ACEI and ARB drugs were excluded (**Table 1** and **Table 2**). Details about the codes of International Statistical Classification of Diseases for identifying prior comorbidities and the items for ACEI/ARB drugs are provided in the **Supplementary Appendix**.

### Statistical analysis and primary outcomes

Primary outcomes are lung cancer and all-cause mortality. Patients were followed up from first ACEI/ARB drug prescription (baseline date) to end points of lung cancer diagnosis. Those without ACEI or ARB drug prescription were followed after hospital admission to study period end (2020/08/31). Follow up ends as the mortality date once the patient died before being diagnosed with lung cancer. Mortality data were obtained from the Hong Kong Death Registry, a population-based official government registry with the registered death records of all Hong Kong citizens. Statistical descriptive statistics are used to summarize characteristics of patients in the lung cancer cohort (**Table 1** and **Table 2**). Continuous variables were presented as median (95% confidence interval [CI] or interquartile range [IQR]) and categorical variables were presented count (%). The Mann-Whitney U test was used to compare continuous variables. The *χ*^2^ test with Yates’ correction was used for 2×2 contingency data, and Pearson’s *χ*^2^ test was used for contingency data for variables with more than two categories.

Using propensity score matching, the ARB inhibitor and ACE groups were then matched at a ratio of 1:3 to form their control groups. Univariate Cox regression was used to identify significant predictors of the primary outcome, with significant variables were further entered into a multivariate Cox regression model with adjustments based on baseline characteristics. Hazard ratios (HRs) with corresponding 95% CIs and P values were reported both for the unmatched and matched cohorts. All significance tests were two-tailed and considered significant if P values were 0.05. Data analyses were performed using RStudio software (Version: 1.1.456) and Python (Version: 3.6).

## Results

### Baseline characteristics

This retrospective population-based cohort was designed to investigate the long-term ACEI/ARB drug exposure effects on lung cancer risk. Initially, 500399 patients first prescribed ACEI/ARB between 1 January 2000 and 31 December 2018 were identified (**Figure 1**). Patients who received both drug classes (n=85563) after the first prescription, or those with a prior diagnosis of lung cancer before initial ACEI/ARB use (n=1127) were excluded. Among the remaining 413709 patients, there were 8452 patients (2.04%) who met the primary lung cancer outcome after a median follow up of 2386 days (IQR: 1141-4067, max: 6500 days).

**Figure 1.**
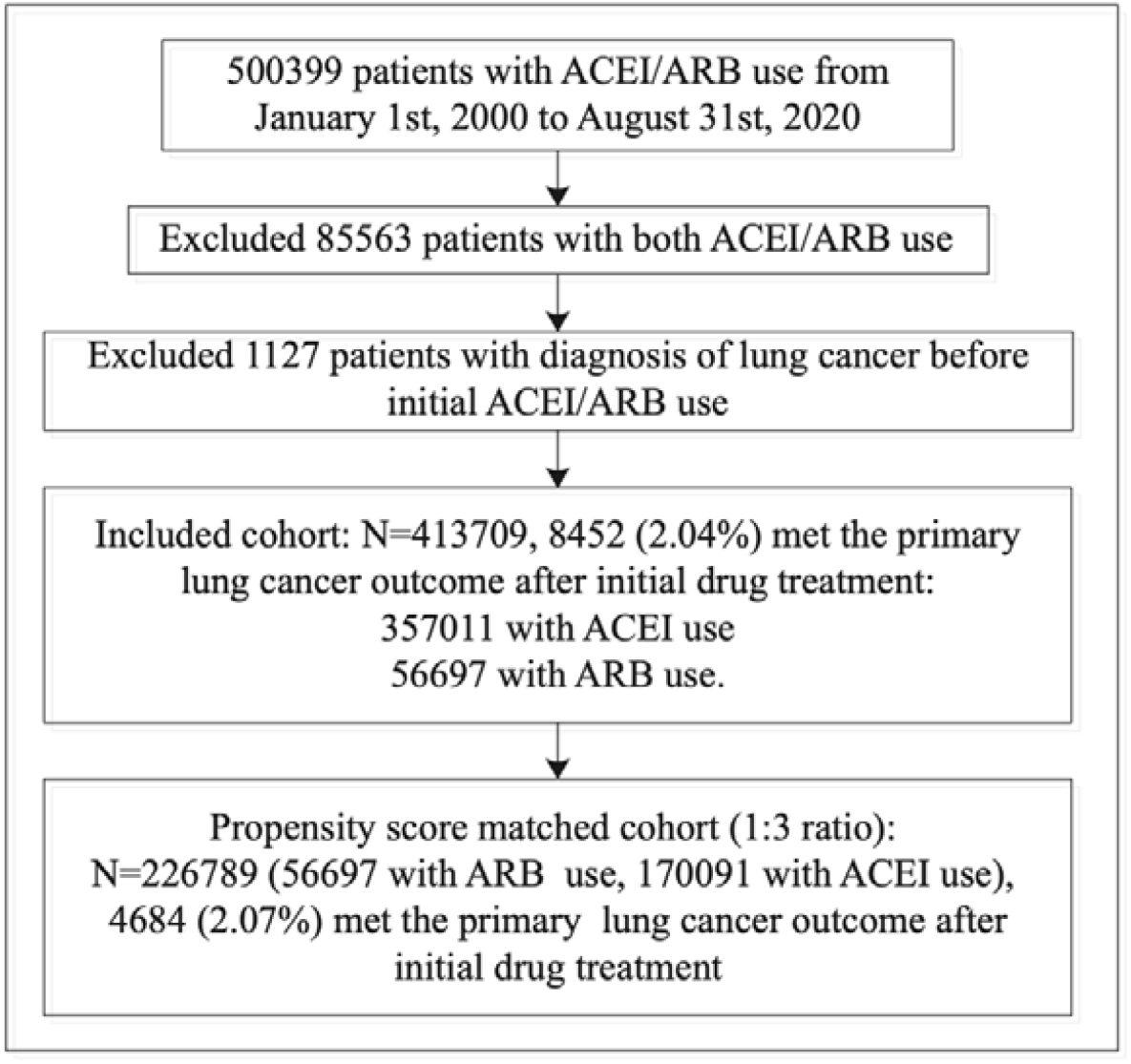
Procedures of data processing.

Of the overall 413708 patients, 357011 patients (86.3%) were prescribed with only ACEI drugs and 56697 patients (13.7%) received treatment with only ARB. On follow-up, the incidence of lung cancer in patients with ACEI use was higher than that with ARB use before propensity score matching (2.16% v.s. 1.29%, P value<0.0001). Kaplan-Meier survival curves is presented in **Figure 2**, and the curves stratified by ACEI or ARB use is provided in **Figure 3**. The comparisons of the clinical baseline characteristics of patients with/without lung cancer were provided in **Supplementary Table 3**.

**Table 3.**
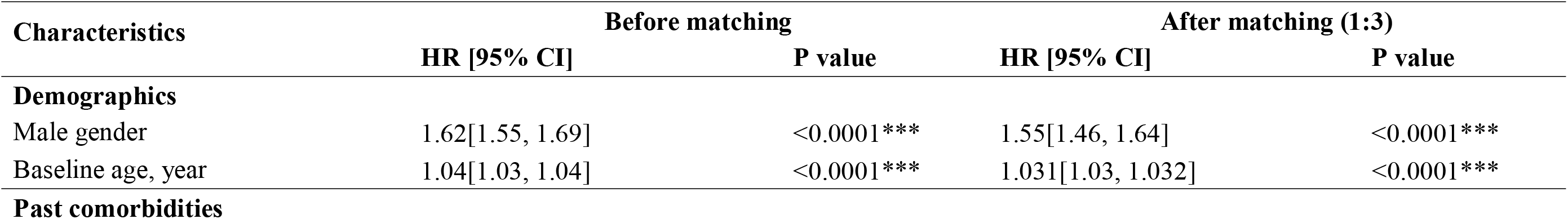
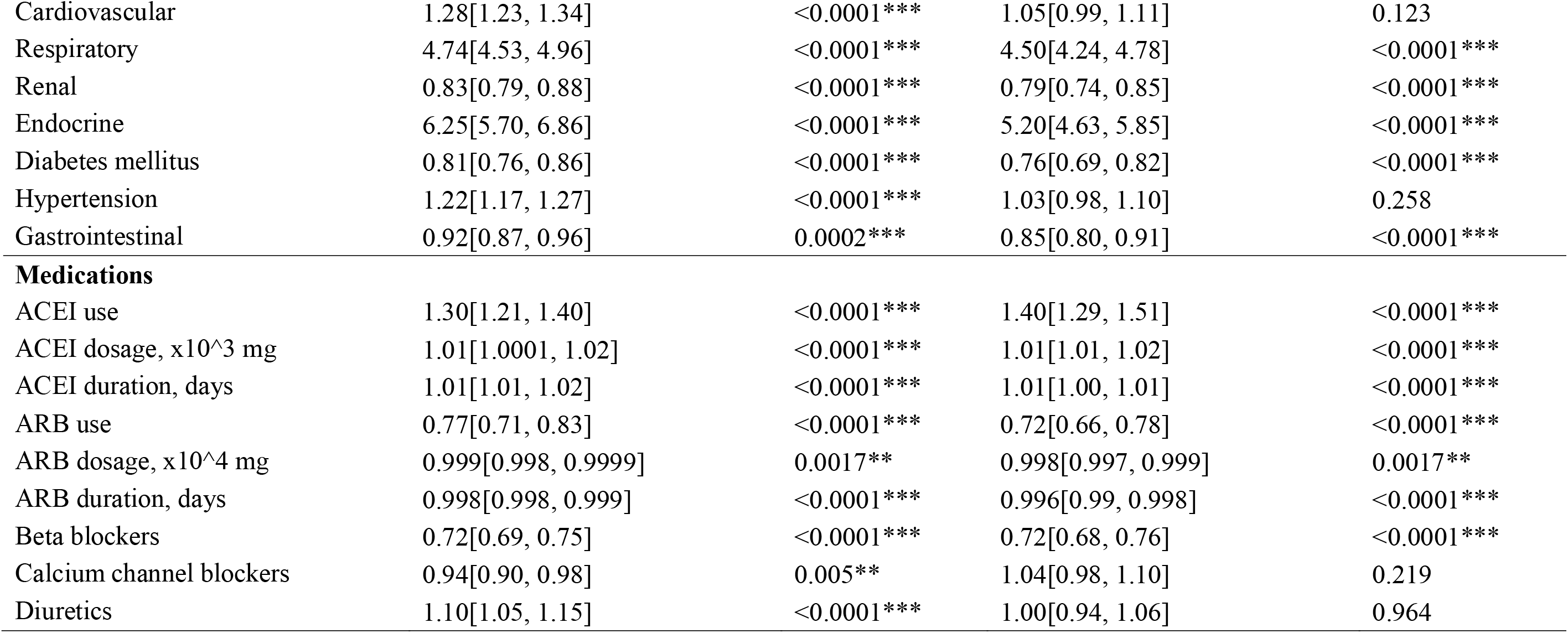
Univariate predictors of incident lung cancer from Cox regression before and after propensity score matching. * for p≤ 0.05, ** for p ≤ 0.01, *** for p ≤ 0.001

| Characteristics | Before matching |  | After matching (1:3) |  |
| --- | --- | --- | --- | --- |
|  | HR [95% CI] | P value | HR [95% CI] | P value |
| **Demographics** |  |  |  |  |
| Male gender | 1.62[1.55, 1.69] | <0.0001*** | 1.55[1.46, 1.64] | <0.0001*** |
| Baseline age, year | 1.04[1.03, 1.04] | <0.0001*** | 1.031[1.03, 1.032] | <0.0001*** |
| **Past comorbidities** |  |  |  |  |
| Cardiovascular | 1.28[1.23, 1.34] | <0.0001*** | 1.05[0.99, 1.11] | 0.123 |
| Respiratory | 4.74[4.53, 4.96] | <0.0001*** | 4.50[4.24, 4.78] | <0.0001*** |
| Renal | 0.83[0.79, 0.88] | <0.0001*** | 0.79[0.74, 0.85] | <0.0001*** |
| Endocrine | 6.25[5.70, 6.86] | <0.0001*** | 5.20[4.63, 5.85] | <0.0001*** |
| Diabetes mellitus | 0.81[0.76, 0.86] | <0.0001*** | 0.76[0.69, 0.82] | <0.0001*** |
| Hypertension | 1.22[1.17, 1.27] | <0.0001*** | 1.03[0.98, 1.10] | 0.258 |
| Gastrointestinal | 0.92[0.87, 0.96] | 0.0002*** | 0.85[0.80, 0.91] | <0.0001*** |
| **Medications** |  |  |  |  |
| ACEI use | 1.30[1.21, 1.40] | <0.0001*** | 1.40[1.29, 1.51] | <0.0001*** |
| ACEI dosage, x10<sup>3</sup> mg | 1.01[1.0001, 1.02] | <0.0001*** | 1.01[1.01, 1.02] | <0.0001*** |
| ACEI duration, days | 1.01[1.01, 1.02] | <0.0001*** | 1.01[1.00, 1.01] | <0.0001*** |
| ARB use | 0.77[0.71, 0.83] | <0.0001*** | 0.72[0.66, 0.78] | <0.0001*** |
| ARB dosage, x10<sup>4</sup> mg | 0.999[0.998, 0.9999] | 0.0017** | 0.998[0.997, 0.999] | 0.0017** |
| ARB duration, days | 0.998[0.998, 0.999] | <0.0001*** | 0.996[0.99, 0.998] | <0.0001*** |
| Beta blockers | 0.72[0.69, 0.75] | <0.0001*** | 0.72[0.68, 0.76] | <0.0001*** |
| Calcium channel blockers | 0.94[0.90, 0.98] | 0.005** | 1.04[0.98, 1.10] | 0.219 |
| Diuretics | 1.10[1.05, 1.15] | <0.0001*** | 1.00[0.94, 1.06] | 0.964 |

**Figure 2.**
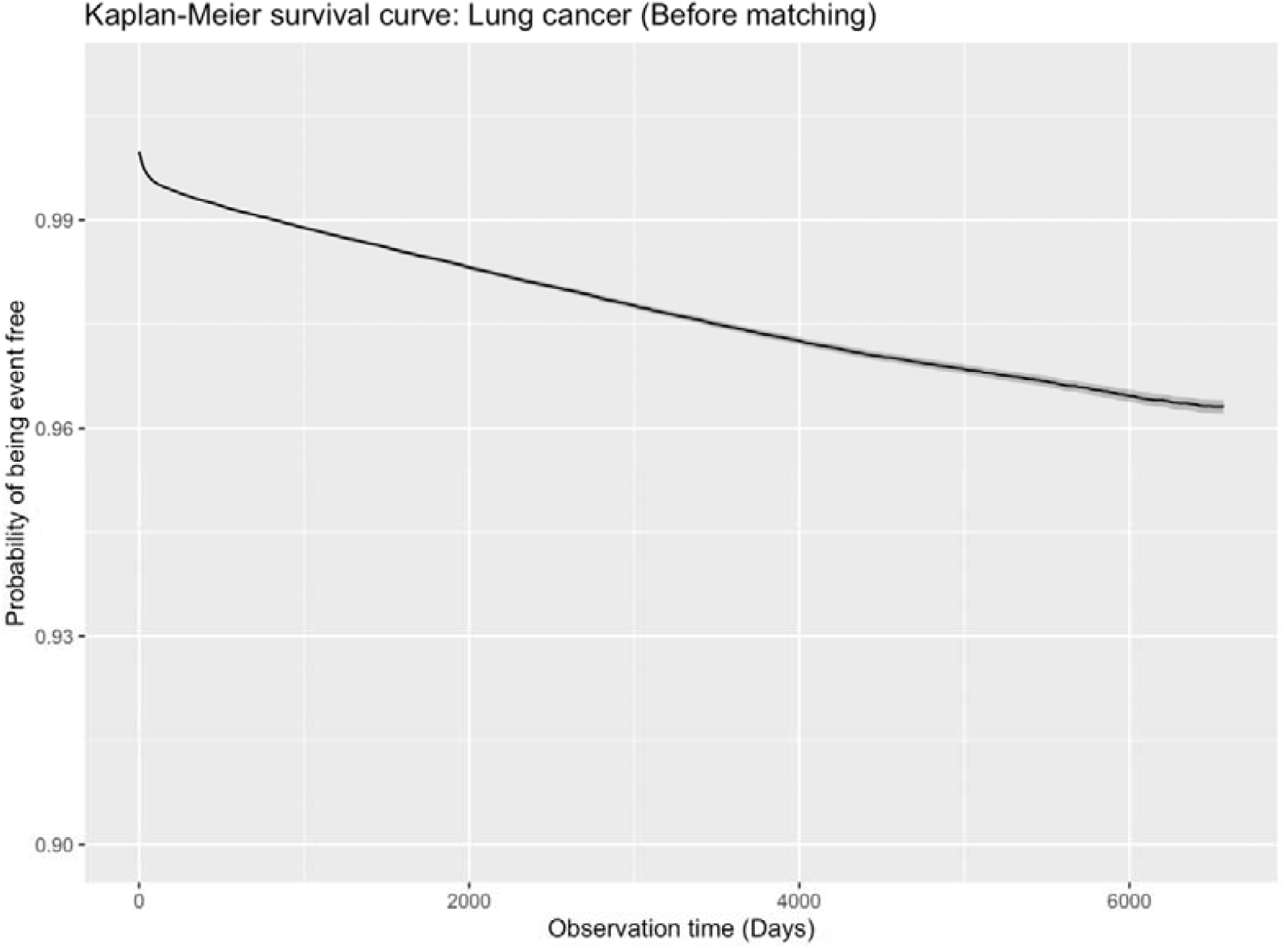
Kaplan-Meier survival curve of lung cancer before propensity score matching.

**Figure 3.**
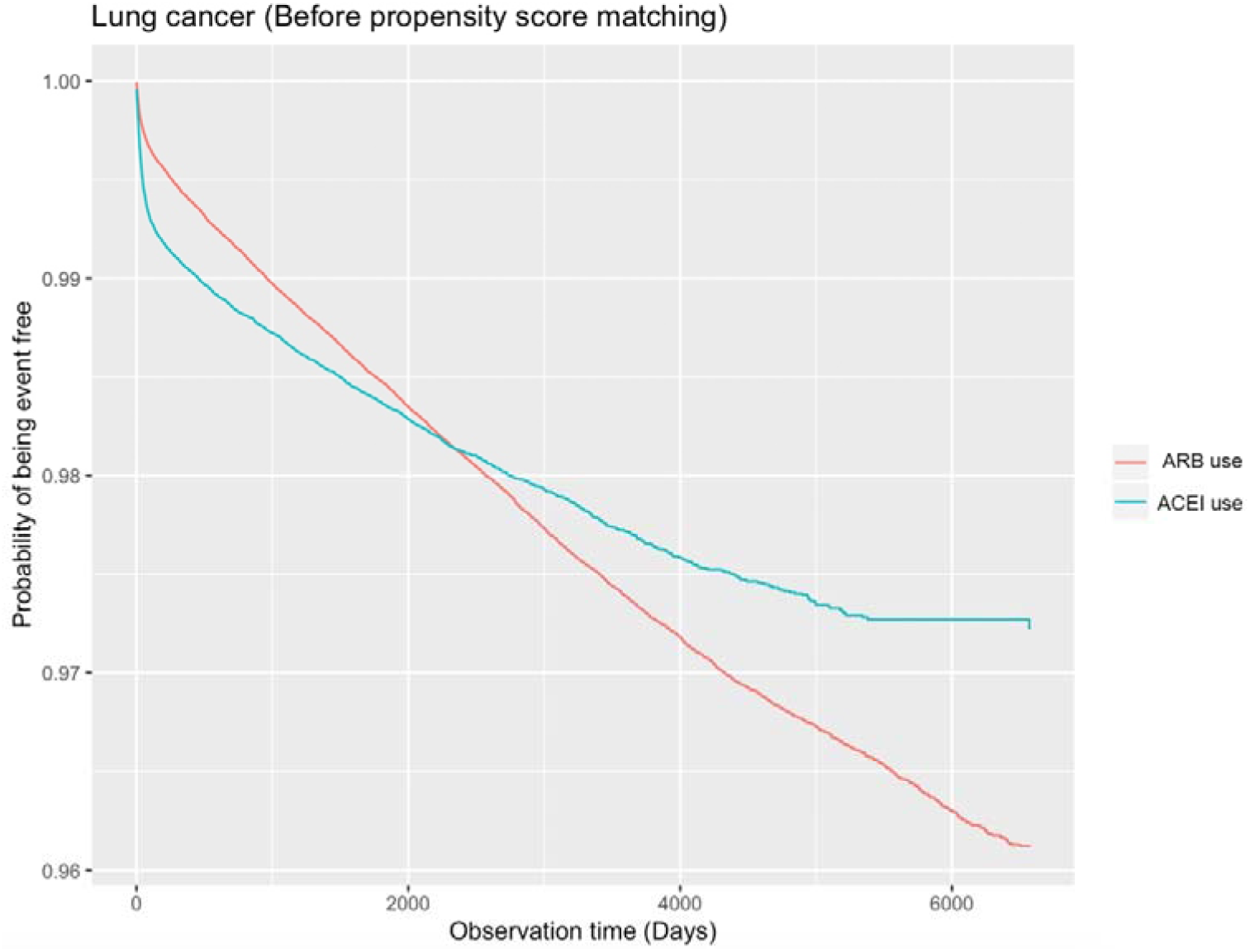
Kaplan-Meier survival curve of lung cancer stratified by ACEI and ARB use before propensity score matching.

### Characteristics after propensity score matching

We conduct propensity score matching approach with 1:3 ratio based on demographics, past comorbidities, and medication classes (**Table 2**). In total 357011 patients (45.86% males) with ACEI use were matched, which is threefold the number of patients with ARB use (45.84%). There are more mortalities of patients with ACEI (50.05% v.s. 13.73%, P value<0.0001) and with longer duration from initial treatment to death (median: 2923 days, IQR: 1452-4498; max: 7477 days v.s. median: 1665 days, IQR: 1074-2742; max: 7304 days, P value<0.0001) than those with ARB use. Higher lung cancer incidence rate of patients with ACEI use was observed (2.32% v.s. 1.29%, P value<0.0001) with longer duration to lung cancer diagnosis (median: 2911 days, IQR: 1436-4488; max: 7477 v.s. median: 1646, IQR: 1060-2717; max: 7304, P value<0.0001). On other significant characteristics differences were observed for the cohort after propensity score matching. Based on the propensity score matched cohort, the Kaplan-Meier survival curve of lung cancer before propensity score matching is presented in **Figure 4**, and the curves stratified by ACEI use and ARB use were provided in **Figure 5**. The comparisons of the clinical baseline characteristics of patients with/without lung cancer for the propensity score matched cohort were provided in **Supplementary Table 4**.

**Figure 4.**
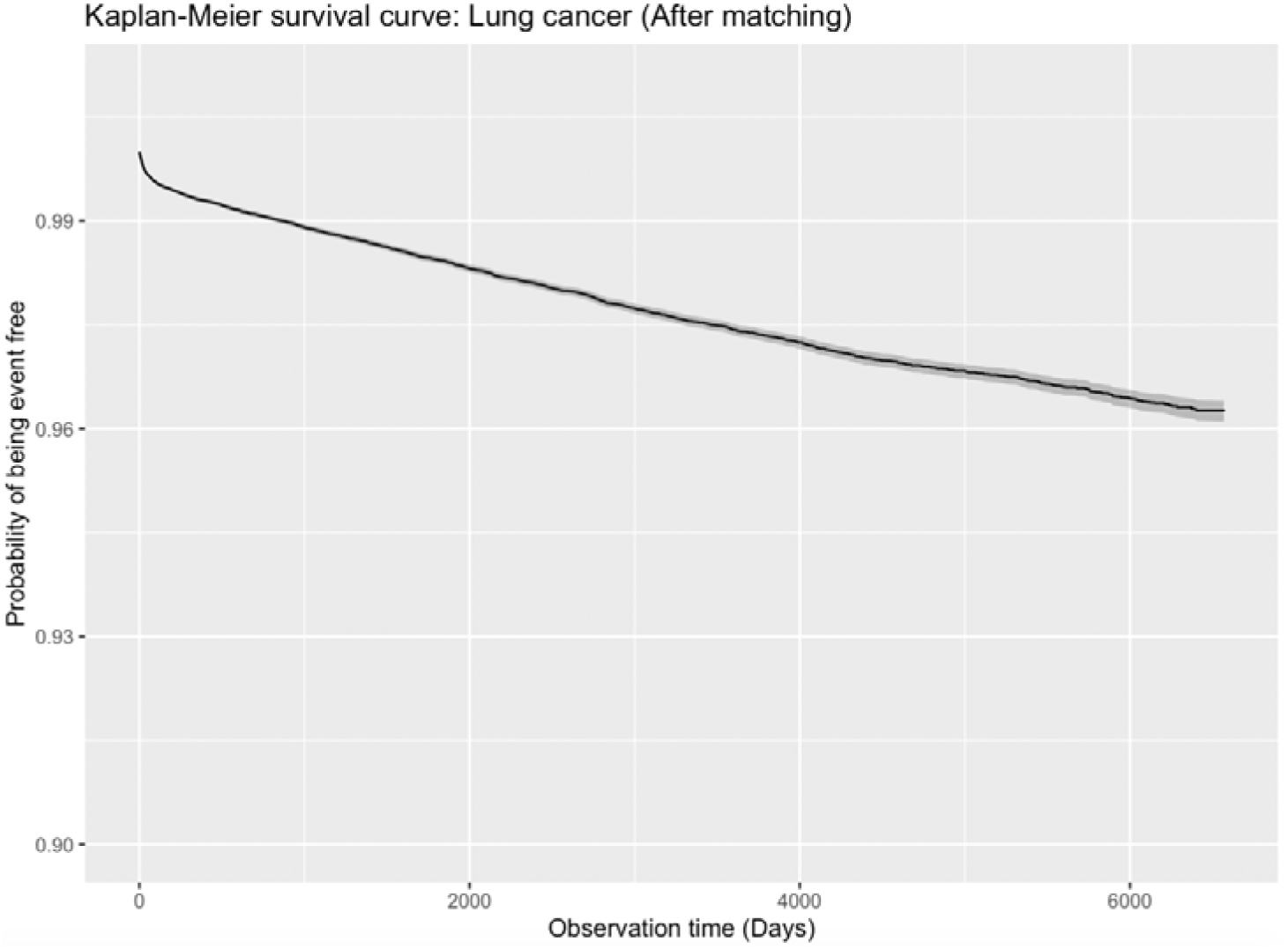
Kaplan-Meier survival curve of lung cancer after propensity score matching.

**Figure 5.**
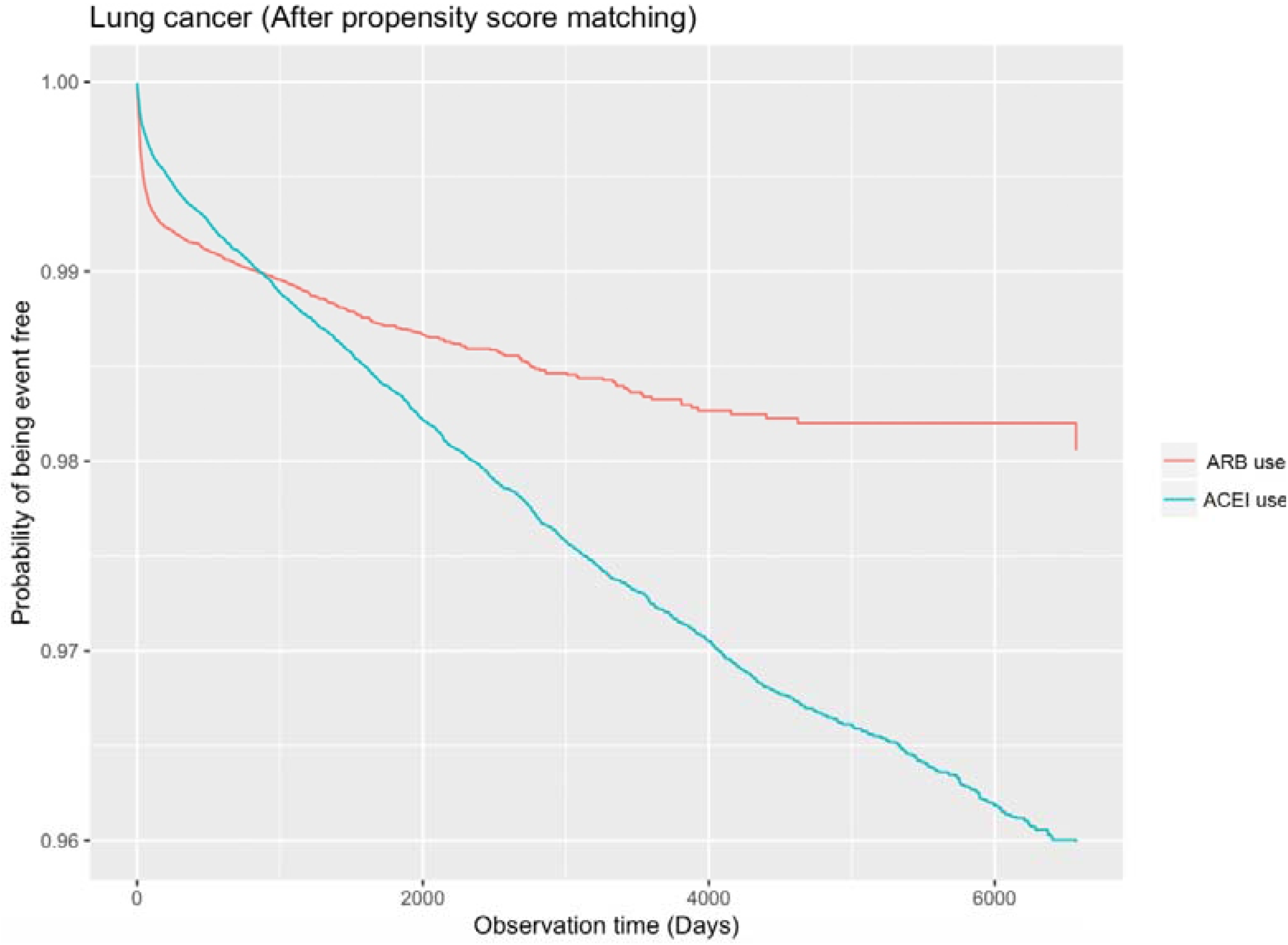
Kaplan-Meier survival curve of lung cancer stratified by ACEI and ARB use after propensity score matching.

**Figure 6.**
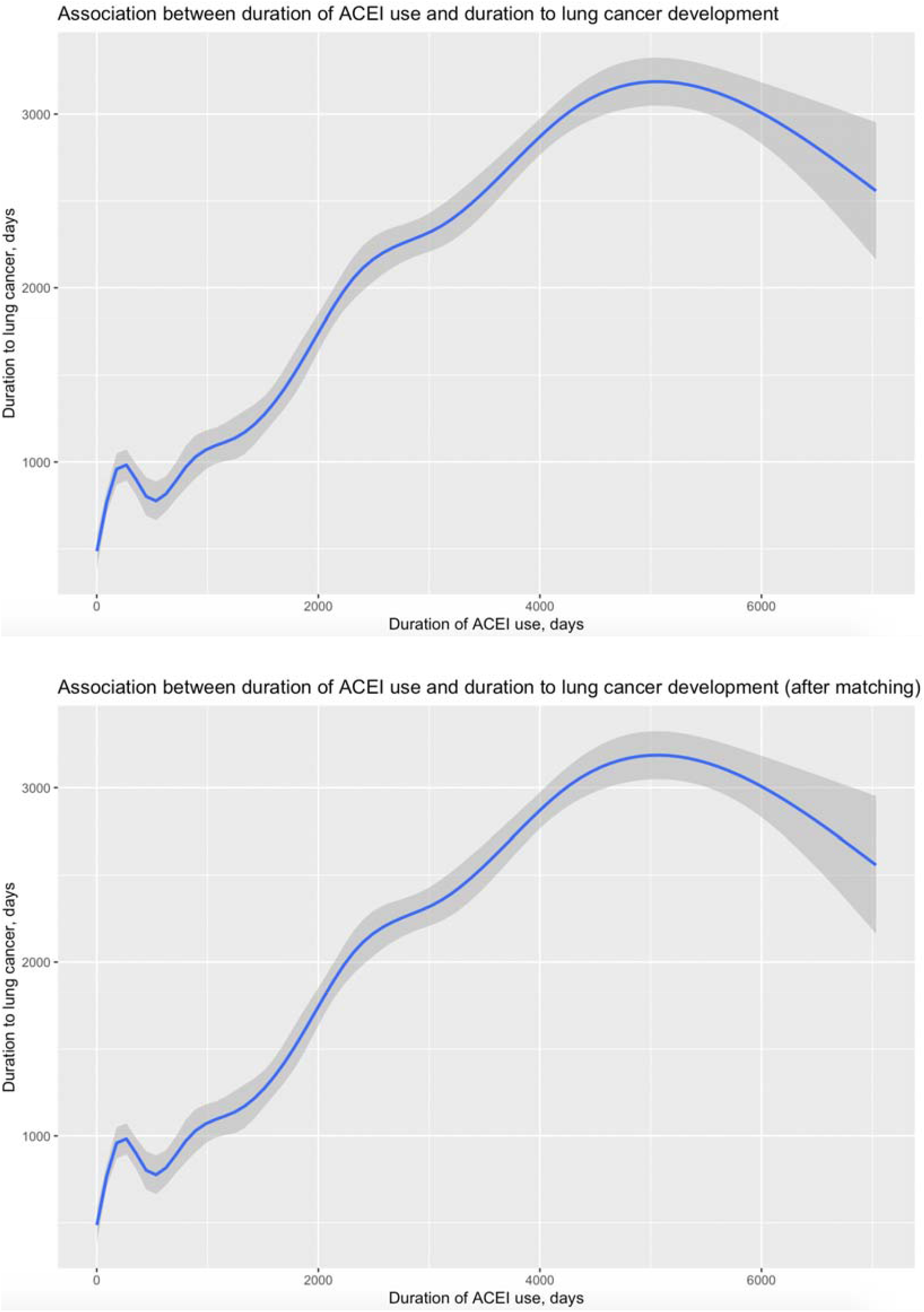
Associations between cumulative ACEI drugs exposure and the duration to lung cancer development before and after propensity score matching.

**Figure 7.**
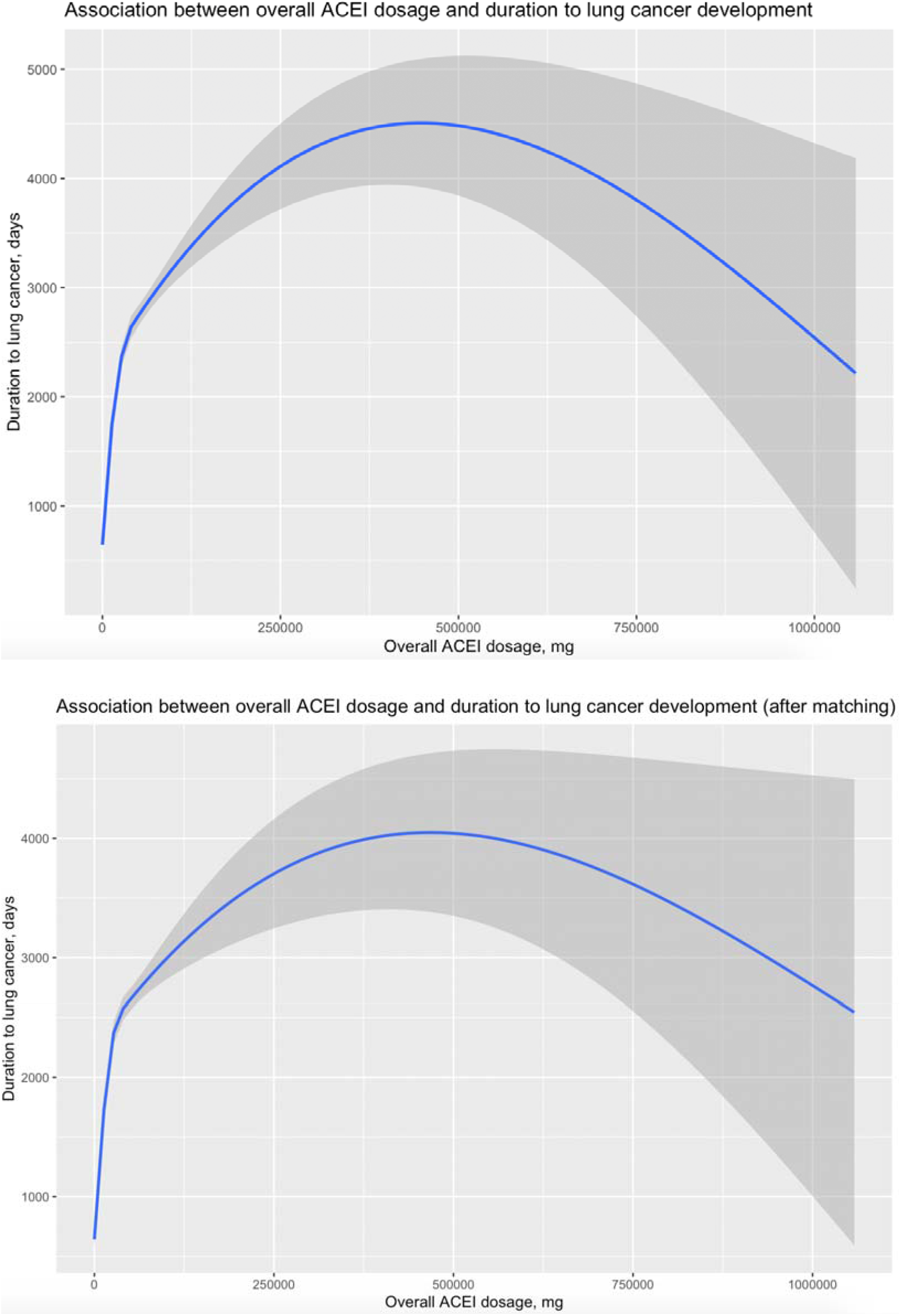
Associations between overall ACEI drugs dosage and the duration to lung cancer development before and after propensity score matching.

**Figure 8.**
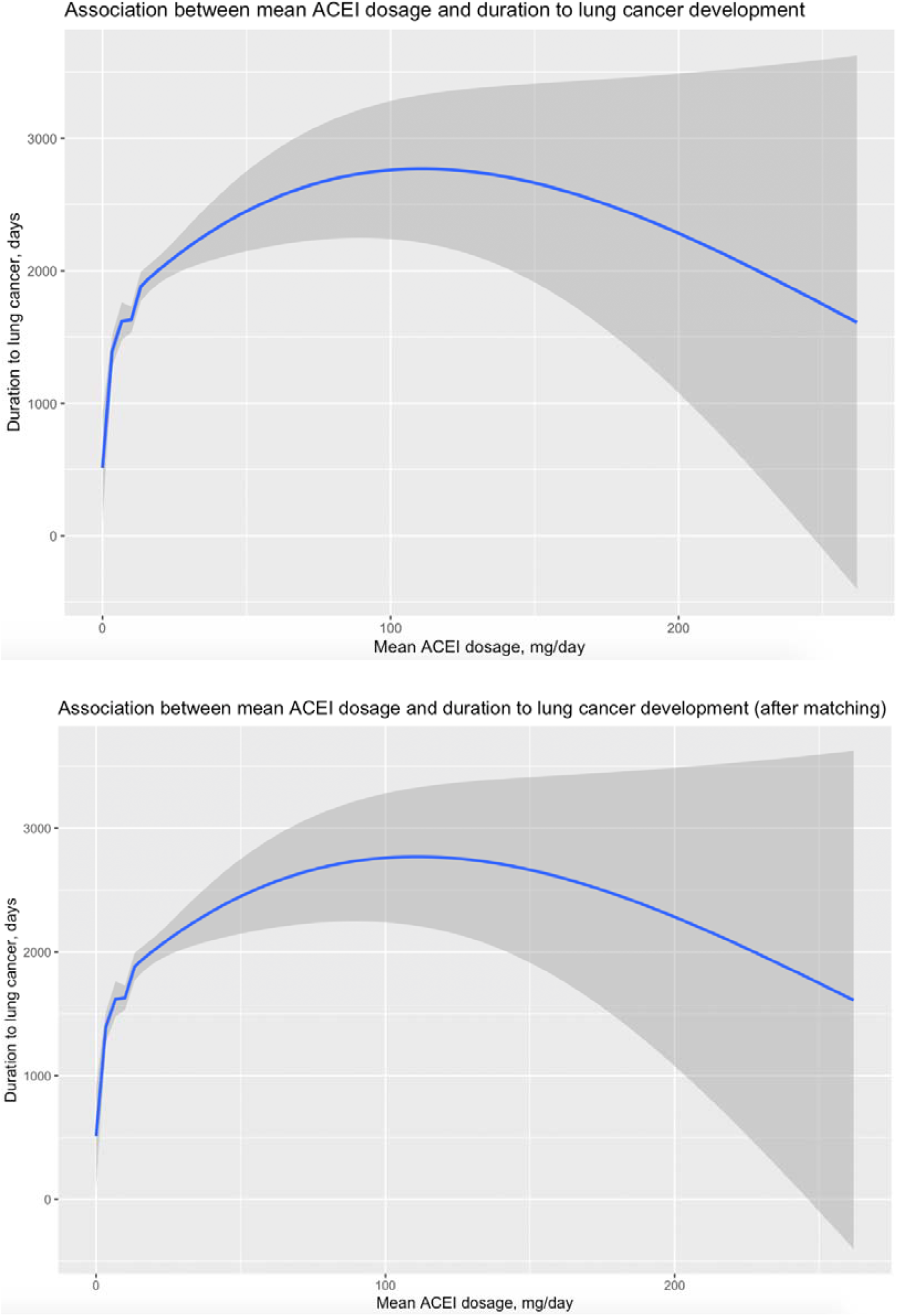
Associations between mean daily ACEI dosage and the duration to lung cancer development before and after propensity score matching.

### Predictors of lung cancer with Cox regression

Univariate Cox regression analysis was conducted to identify the significant predictors associated with lung cancer diagnosis as shown in **Table 3**, including male gender (HR: 1.62, 95% CI: 1.55-1.69, P value<0.0001), older baseline age (HR: 1.04, 95% CI: 1.03-1.04, P value<0.0001); past comorbidities of cardiovascular diseases (HR: 1.28, 95% CI: 1.23-1.34, P value<0.0001), respiratory diseases (HR: 4.74, 95% CI: 4.53-4.96, P value<0.0001), endocrine diseases (HR: 6.25, 95% CI: 5.70-6.86, P value<0.0001), hypertension (HR: 1.22, 95% CI: 1.17-1.27, P value<0.0001); prescription of ACEI drugs (HR: 1.3, 95% CI: 1.21-1.4, P value<0.0001) with larger dosage amount (HR: 1.01, 95% CI: 1.0001, 1.02, P value<0.0001) and longer duration (HR: 1.01, 95% CI: 1.01-1.02, P value<0.0001) and prescriptions of diuretics (HR: 1.1, 95% CI: 1.05-1.15, P value<0.0001). Prescription of ARB drugs was not significantly associated with lung cancer diagnosis (HR<1, P value<0.05).

Significant risk factors associated with lung cancer diagnosis after propensity score matching include male gender (HR: 1.55, 95% CI: 1.46-1.64, P value<0.0001), older baseline age (1HR: 1.031, 95% CI: 1.03-1.032, P value<0.0001); past comorbidities of respiratory diseases (HR: 4.5, 95% CI: 4.24, 4.78, P value<0.0001), endocrine diseases (HR: 5.20, 95% CI: 4.63-5.85, P value<0.0001); prescriptions of ACEI drugs (HR: 1.4, 95% CI: 1.29-1.51, P value<0.0001), with larger amount (HR: 1.01, 95% CI: 1.01-1.02, P value<0.0001) and longer duration (HR: 1.01, 95% CI: 1.00-1.01, P value<0.0001). Prescriptions of ARB drugs and beta blockers doesn’t demonstrate significant effects with lung cancer diagnosis (HR<1, P value<0.05).

The significant predictors before and after propensity score matching were used as input of multivariate Cox regression models respectively for further identification, as shown in **Table 4**. After propensity scoring matching, male gender (HR: 1.79, 95% CI: 1.69-1.9, P value<0.0001), older baseline age (HR: 1.031, 95% CI: 1.03-1.032], P value<0.0001); prior comorbidities of respiratory diseases (HR: 5.04, 95% CI: 4.74-5.36, P value<0.0001) and endocrine diseases (HR: 4.54, 95% CI: 4.03-5.11, P value<0.0001); prescription of ACEI drugs (HR: 1.48, 95% CI: 1.36-1.6, P value<0.0001) with longer duration (HR: 1.01, 95% CI: 1.00-1.01, P value=0.0009). Prescriptions of ARB drugs were not significantly associated with lung cancer diagnosis (HR<1, P value<0.05).

**Table 4.**
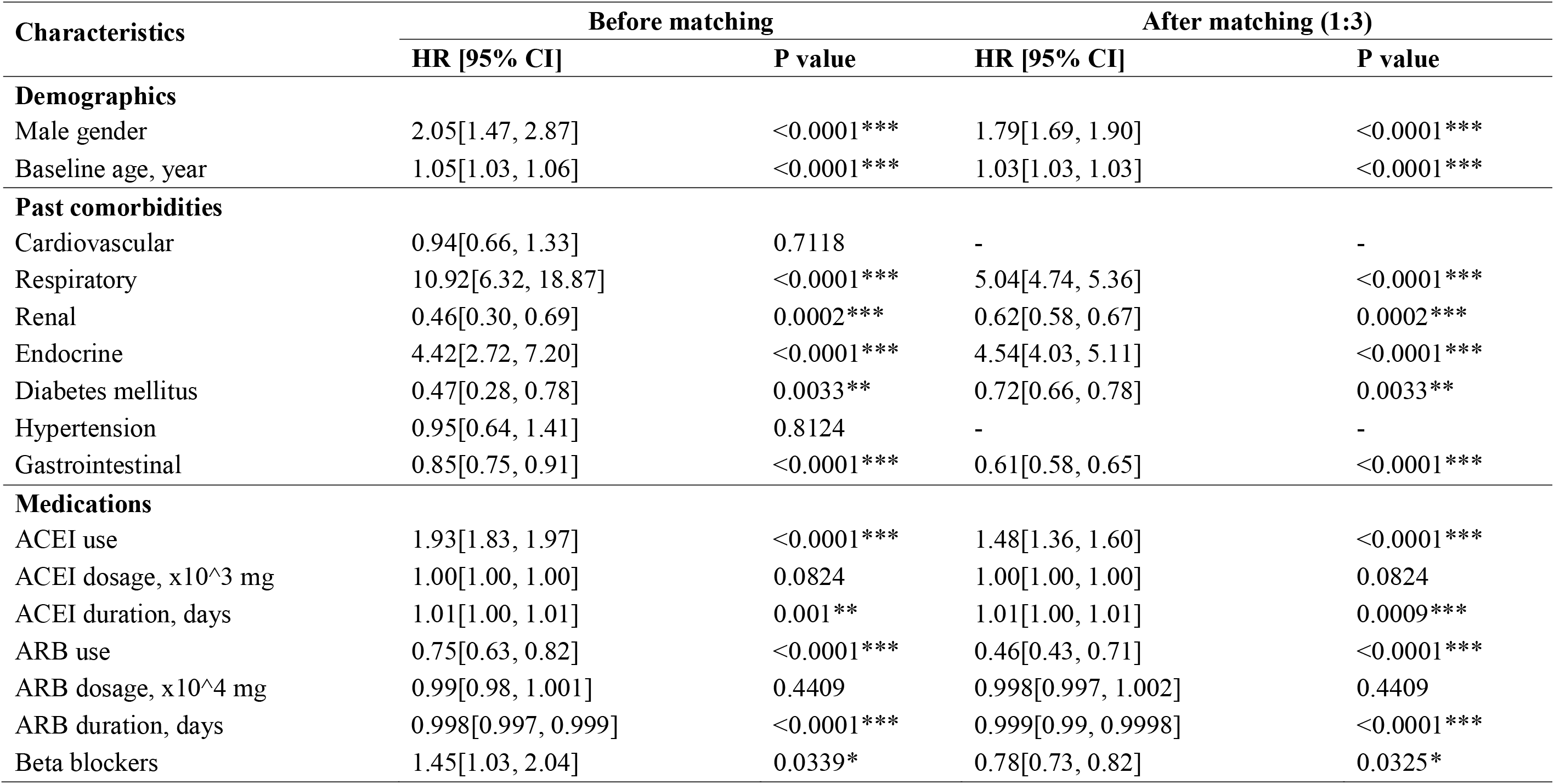
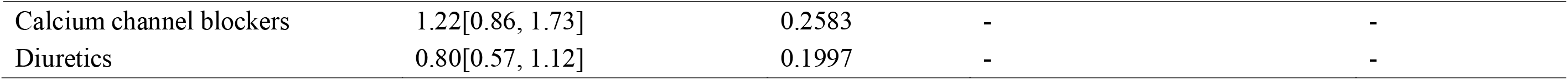
Multivariate Cox analysis of significant predictors of incident lung cancer. * for p≤ 0.05, ** for p ≤ 0.01, *** for p ≤ 0.001

| Characteristics | Before matching |  | After matching (1:3) |  |
| --- | --- | --- | --- | --- |
|  | HR [95% CI] | P value | HR [95% CI] | P value |
| **Demographics** |  |  |  |  |
| Male gender | 2.05[1.47, 2.87] | <0.0001*** | 1.79[1.69, 1.90] | <0.0001*** |
| Baseline age, year | 1.05[1.03, 1.06] | <0.0001*** | 1.03[1.03, 1.03] | <0.0001*** |
| **Past comorbidities** |  |  |  |  |
| Cardiovascular | 0.94[0.66, 1.33] | 0.7118 | - | - |
| Respiratory | 10.92[6.32, 18.87] | <0.0001*** | 5.04[4.74, 5.36] | <0.0001*** |
| Renal | 0.46[0.30, 0.69] | 0.0002*** | 0.62[0.58, 0.67] | 0.0002*** |
| Endocrine | 4.42[2.72, 7.20] | <0.0001*** | 4.54[4.03, 5.11] | <0.0001*** |
| Diabetes mellitus | 0.47[0.28, 0.78] | 0.0033** | 0.72[0.66, 0.78] | 0.0033** |
| Hypertension | 0.95[0.64, 1.41] | 0.8124 | - | - |
| Gastrointestinal | 0.85[0.75, 0.91] | <0.0001*** | 0.61[0.58, 0.65] | <0.0001*** |
| **Medications** |  |  |  |  |
| ACEI use | 1.93[1.83, 1.97] | <0.0001*** | 1.48[1.36, 1.60] | <0.0001*** |
| ACEI dosage, $\times 10^3$ mg | 1.00[1.00, 1.00] | 0.0824 | 1.00[1.00, 1.00] | 0.0824 |
| ACEI duration, days | 1.01[1.00, 1.01] | 0.001** | 1.01[1.00, 1.01] | 0.0009*** |
| ARB use | 0.75[0.63, 0.82] | <0.0001*** | 0.46[0.43, 0.71] | <0.0001*** |
| ARB dosage, $\times 10^4$ mg | 0.99[0.98, 1.001] | 0.4409 | 0.998[0.997, 1.002] | 0.4409 |
| ARB duration, days | 0.998[0.997, 0.999] | <0.0001*** | 0.999[0.99, 0.9998] | <0.0001*** |
| Beta blockers | 1.45[1.03, 2.04] | 0.0339* | 0.78[0.73, 0.82] | 0.0325* |
| Calcium channel blockers | 1.22[0.86, 1.73] | 0.2583 | - | - |
| Diuretics | 0.80[0.57, 1.12] | 0.1997 | - | - |

## Discussions

In this territory-wide population-based study, the use of ACEIs was associated with increased risk of both lung and gastrointestinal cancer compared with ARB use at all time points. This study features the largest cohort groups of patients prescribed with ACEI (500,997) and ARB (120,917) to date. Data was collected from a territory-wide clinical database involved 42 hospitals, with the longest follow-up period (1 January 2001 and 31 December 2018) to date. ACEI use was found to increase both lung and gastrointestinal cancer risk at all time-points, including within 1 year of ACEI initiation. Moreover, the incidence rate increased with time since initiation.

Preclinical studies have investigated the association between ACEIs and carcinogenesis. Angiotensin-converting enzyme metabolizes neuropeptides substance P and bradykinin, which therefore accumulate with ACEI use. Studies have shown ACE expression in the lung, gastrointestinal tract, and urogenital system. Substance P has been shown to facilitate cancer proliferation and progression, as well as promote angiogenesis. Bradykinin has been documented as a pluripotent factor for cancer growth by direct stimulation of cancer growth, inducing vascular endothelial growth factor release, and assisting tumor invasion and migration through metalloproteases [11, 12]. Lung cancer tissue is known to express substance P and bradykinin receptors. As such, ACEI use may induce carcinogenesis through such neuropeptide mechanisms in tissues that express ACE.

The use of ACEIs has been associated with bradykinin accumulation in the lungs [13], which in turn may stimulate lung cancer growth [14]. However, epidemiological data on the association between the use of ACEIs and cancer risk at the population level are conflicting. Several meta-analyss of randomized controlled trials have shown no association, but are limited by insufficient power to assess cancer outcomes [9, 10]. In terms of observational studies, many have short durations of follow-up of less than 5 years, and such are not able to assess long-term outcomes such as lifetime cancer risk [15-19]. A comparative population-based cohort study of ACEI and ARB use in the United States found no difference in incident lung cancer in ACEI versus ARB initiators (HR = 0.99, 95% CI= [0.84, 1.16]) but was limited by a short follow up of 5 years [20]. In comparison, a recent population-based cohort study in the United Kingdom reported a 14% increase in lung cancer incidence with long-term use of ACEIs compared to ARBs [7]. This UK study had a relatively moderately long follow-up period of 6.4 years with a large cohort size of more than 990 000 patients. This study also found a higher lung cancer risk in ACEI users compared to non-users. By contrast, ARB users showed significantly lower lung cancer risk. Our study results agree with those from the United Kingdom and Taiwan cohorts, which would suggest generalizability to the general population.

Moreover, studies on ACEI use and gastrointestinal cancer have been comparatively sparse. A nested case-control study of 10 000 individuals was conducted using the General Practitioners’ Research Database aged 40–84 years between 1994 and 2001 [21]. This reported a lower risk of esophageal adenocarcinoma by 29%, but no significant difference in squamous-cell carcinoma risk compared with nonusers. Similarly, no clear association between the use of ACE inhibitors and the risk of gastric cancer was observed. In another study, long-term lisinopril use was associated with a lower incidence of advanced adenomatous colon polyps [22]. Other studies, in comparison, found no protective effects of ACEIs for colorectal cancer [23]. A meta-analysis of randomized controlled trials found that ACEI therapy did not have an effect on gastrointestinal cancer risk [24]. Our study nevertheless adds to the literature by reporting a significantly higher incidence of GI cancers in ACEI users compared to ARB users.

### Strengths and limitations

This study included data from the administrative database, which includes all patient data from every public hospital and clinic in Hong Kong serving patients from all geographical and socioeconomic status. As such, the cohort is likely an accurate representation of the Hong Kong population. Moreover, with a study cohort of more than 600 000 patients, this is the largest study of ACEI or ARB users investigating the association of ACEI/ARB and cancer incidence. The long follow-up period of the study also assists in the investigation of long-term outcomes such as cancer incidence.

The limitations of this study include the absence of data on some potential confounders such as smoking, diet, and other recreational drug use. Information such as smoking habits and pack years are not recorded in the CDARS. Studies have shown the strong association between smoking and duration of smoking with certain types of lung cancers, as well as the relationship between diet and colorectal cancer. However, the use of the ARB study cohort as a control and the significant cohort size should limit the impact of these confounding factors on the study findings. Propensity matching could be used to further account for these limitations. Secondly, patients may have independently influenced their exposure to ACEI/ARB use by non-compliance, resulting in misclassification. However, the rates of non-compliance should be minimal between the ACEI and ARB groups, and thus should not affect the comparative analysis of incident cancer.

## Conclusions

ACEI use was associated with increased risk of lung cancer compared with ARB use at all time points. Additionally, incidence risk increases with the duration of exposure. Future studies should validate our findings in other cohorts.

## Supporting information

STROBE checklist

