## Supplementary Appendix for "Angiotensin converting enzyme inhibitors and risk of lung cancer: a population-based cohort study"

**Supplementary Table 1. Codes for identifying lung cancer**

| **Lung Cancer Code (ICD9)** | **Lung Cancer Description (ICD9)** |
| --- | --- |
| **160 Malignant neoplasm of nasal cavities, middle ear, and accessory sinuses** | |
| 160.0 | Mal neo nasal cavities |
| 160.1 | Malig neo middle ear |
| 160.2 | Mal neo maxillary sinus |
| 160.3 | Mal neo ethmoidal sinus |
| 160.4 | Malig neo frontal sinus |
| 160.5 | Mal neo sphenoid sinus |
| 160.8 | Mal neo access sinus NEC |
| 160.9 | Mal neo access sinus NOS |
| **161 Malignant neoplasm of larynx** | |
| 161.0 | Malignant neo glottis |
| 161.1 | Malig neo supraglottis |
| 161.2 | Malig neo subglottis |
| 161.3 | Mal neo cartilage larynx |
| 161.8 | Malignant neo larynx NEC |
| 161.9 | Malignant neo larynx NOS |
| **162 Malignant neoplasm of trachea, bronchus, and lung** | |
| 162.0 | Malignant neo trachea |
| 162.2 | Malig neo main bronchus |
| 162.3 | Mal neo upper lobe lung |
| 162.4 | Mal neo middle lobe lung |
| 162.5 | Mal neo lower lobe lung |
| 162.8 | Mal neo bronch/lung NEC |
| 162.9 | Mal neo bronch/lung NOS |
| **163 Malignant neoplasm of pleura** | |
| 163.0 | Mal neo parietal pleura |
| 163.1 | Mal neo visceral pleura |
| 163.8 | Malig neopl pleura NEC |
| 163.9 | Malig neopl pleura NOS |
| **164 Malignant neoplasm of thymus, heart, and mediastinum** | |
| 164.0 | Malignant neopl thymus |
| 164.1 | Malignant neopl heart |
| 164.2 | Mal neo ant mediastinum |
| 164.3 | Mal neo post mediastinum |
| 164.8 | Mal neo mediastinum NEC |
| 164.9 | Mal neo mediastinum NOS |
| **165 Malignant neoplasm of other and ill-defined sites within the respiratory system and intrathoracic organs** | |
| 165.0 | Mal neo upper resp NOS |
| 165.8 | Mal neo thorax/resp NEC |
| 165.9 | Mal neo resp system NOS |
| 197.0 | Lung cancer spread from another organ |
| 197.7 | Lung cancer spreads to liver |
| 198.3 | Lung cancer spreads to brain |
| 198.5 | Lung cancer spreads to bone |
| 198.7 | Lung cancer spreads to adrenal glands |
| 212.3 | Nonmalignant neoplasms of the lung - uncertain behavior |
| 235.7 | Nonmalignant neoplasms of the lung - benign |
| 239.1 | Nonmalignant neoplasms of the lung - unspecified nature |

**Supplementary Table 2. ACEI and ARB drugs**

**ACEI drugs**

LISINOPRIL, MALEATE, RAMIPRIL, CAPTOPRIL, ALAPRIL, ENALAPRIL, ACERTIL, ARGININE, CAPTOPRI, CAPTOPRILTABLET, CILAZAPRIL, DOCUMED:, EROOM, FOSINOPRIL, PERINDOPRIL, PERINDOPRILTERTBUTYLAMINE, PRIVATE, QUINAPRIL

**ARB drugs**

AZILSARTAN, CANDESARTAN, CILEXETIL, CO-DIOVAN, DIOVAN, ENTRESTO, IRBESARTAN, LOSARTAN, MICARDIS, SPARSENTAN/IRBESARTAN, TELMISARTAN, VALSARTAN

**Supplementary Table 3. Clinical baseline characteristics of lung cancer patients and non-lung cancer patients**

* for p≤ 0.05, ** for p ≤ 0.01, *** for p ≤ 0.001

| **Characteristics** | **Lung cancer (N=**10797**)** | **No lung cancer (N=**528798**)** | | | **P value** |
| --- | --- | --- | --- | --- | --- |
| Mortality | 9365(86.73%) | 245715(46.46%) | | | <0.0001*** |
| Duration to mortality, days | 1594(586-3063);6357;n=10797 | 2420(1169-4096);5529;n=528798 | | | <0.0001*** |
| **Demographics** |  | |  | |  |
| Male gender | 7041(65.21%) | 280310(53.00%) | | | <0.0001*** |
| Baseline age at drug prescription, year | 73.2(66.2-79.3);100.9;n=10797 | 71.1(56.0-79.8);116.3;n=528798 | | | <0.0001*** |
| **Past comorbidities** |  | |  | |  |
| Cardiovascular | 5087(47.11%) | 187685(35.49%) | | | <0.0001*** |
| Respiratory | 7573(70.13%) | 140116(26.49%) | | | <0.0001*** |
| Renal | 2083(19.29%) | 100192(18.94%) | | | 0.4625 |
| Endocrine | 867(8.03%) | 3878(0.73%) | | | <0.0001*** |
| Diabetes mellitus | 1686(15.61%) | 84262(15.93%) | | | 0.4521 |
| Hypertension | 6167(57.11%) | 235972(44.62%) | | | <0.0001*** |
| Gastrointestinal | 3474(32.17%) | 153741(29.07%) | | | <0.0001*** |
| **Medications** |  | | |  |  |
| ACEI use | 7719(71.49%) | 349292(66.05%) | | | <0.0001*** |
| ACEI dosage, x10^3 mg | 3.82(0.61-15.5);241.02;n=7719 | 5.99 (0.87-24.7);434.9;n=349291 | | | <0.0001*** |
| ACEI duration, days | 866.0(156.5-2336.0);20618.0;n=6382 | 1285.0(219.0-3265.0);26666.0;n=299887 | | | <0.0001*** |
| ARB use | 733(6.78%) | 55964(10.58%) | | | <0.0001*** |
| ARB dosage, x10^4 mg | 4.75(1.03-10.98);71.2;n=54 | 7.9 (1.98-19.23);427.3;n=8011 | | | 0.0063** |
| ARB duration, days | 891.0(233.5-1588.0);5021.0;n=47 | 1701.0(826.0-2890.0);12870.0;n=7749 | | | <0.0001*** |
| Beta blockers | 4272(39.56%) | 244376(46.21%) | | | <0.0001*** |
| Calcium channel blockers | 5981(55.39%) | 299618(56.66%) | | | 0.1679 |
| Diuretics | 4241(39.27%) | 208595(39.44%) | | | 0.8232 |

**Supplementary Table 4. Clinical baseline characteristics of lung cancer patients and non-lung cancer patients after propensity score matching**

* for p≤ 0.05, ** for p ≤ 0.01, *** for p ≤ 0.001

| **Characteristics** | **Lung cancer (N=**10797**)** | **No lung cancer (N=**528798**)** | | | **P value** |
| --- | --- | --- | --- | --- | --- |
| Mortality | 3800(81.12%) | 89120(40.12%) | | | <0.0001*** |
| Duration to mortality, days | 1774(781.5-3203.5);7257;n=4684 | 2516(1269-4189);7177;n=222104 | | | <0.0001*** |
| **Demographics** |  | |  | |  |
| Male gender | 2633(56.21%) | 101368(45.63%) | | | <0.0001*** |
| Baseline age at drug prescription, year | 72.4(65.1-79.7);100.9;n=4684 | 70.5 (60-80.1);110.2;n=222104 | | | <0.0001*** |
| **Past comorbidities** |  | |  | |  |
| Cardiovascular | 1960(41.84%) | 89824(40.44%) | | | 0.215 |
| Respiratory | 3031(64.70%) | 65723(29.59%) | | | <0.0001*** |
| Renal | 838(17.89%) | 47189(21.24%) | | | <0.0001*** |
| Endocrine | 298(6.36%) | 2602(1.17%) | | | <0.0001*** |
| Diabetes mellitus | 614(13.10%) | 35126(15.81%) | | | <0.0001*** |
| Hypertension | 2794(59.64%) | 126710(57.04%) | | | 0.0669 |
| Gastrointestinal | 1542(32.92%) | 79690(35.87%) | | | 0.0039** |
| **Medications** |  | | |  |  |
| ACEI use | 3951(84.35%) | 166140(74.80%) | | | <0.0001*** |
| ACEI dosage, x10^3 mg | 4.1 (0.6-15.8);320.7;n=3951 | 6.3(0.9-25.5);371.2;n=166139 | | | <0.0001*** |
| ACEI duration, days | 898(175-2368);20618;n=3339 | 1310(230-3327);6408;n=143524 | | | <0.0001*** |
| ARB use | 733(15.64%) | 55964(25.19%) | | | <0.0001*** |
| ARB dosage, x10^4 mg | 4.8 (1.03-11.0);71.2;n=54 | 7.9 (1.98-19.23);127.3;n=8011 | | | 0.0065** |
| ARB duration, days | 891(233.5-1588);5021;n=47 | 1701(826-2890);4870;n=7749 | | | <0.0001*** |
| Beta blockers | 1854(39.58%) | 103308(46.51%) | | | <0.0001*** |
| Calcium channel blockers | 2843(60.69%) | 134814(60.69%) | | | 0.9918 |
| Diuretics | 1533(32.72%) | 77047(34.68%) | | | 0.0519 |
